# Vaccine effectiveness and duration of protection against symptomatic and severe Covid-19 during the first year of vaccination in France

**DOI:** 10.1101/2022.02.17.22270791

**Authors:** Milena Suarez Castillo, Hamid Khaoua, Noémie Courtejoie

## Abstract

**Background:** SARS-CoV-2 continues to spread despite fast vaccine rollout, which could be attributed to waning immunity or to a reduced protection against some variants. A thorough characterization of vaccine protection and its duration in time is needed to inform vaccination policies and enhance public trust.

**Methods:** We matched three national databases with exhaustive information on screening, vaccination and hospitalizations in France over the year 2021. We performed a two-step analysis to estimate vaccine effectiveness against severe forms of Covid-19 in people aged 50 years or over, combining: *(i)* a test-negative case–control design to assess vaccine effectiveness against symptomatic infections; and *(ii)* a survival analysis to assess the additional protection against severe outcomes (hospitalizations and inpatient deaths) in infected individuals.

**Results:** We found a high vaccine effectiveness in people aged 50 years or more, reaching 82% against symptomatic infections and 94% against severe outcomes, after a full vaccination scheme.

Vaccine effectiveness against symptomatic infections strongly decreased over time, dropping to 53% after six months, but remained high against severe forms (90% after six months). The booster dose allowed restoring high protection levels. Vaccine protection and its evolution in time, showed little difference against the variants that circulated prior to December 2021 in France, including the Delta variant.

**Conclusion:** Though vaccine immunity decreases over time, vaccination remains crucial to provide individual protection against severe diseases. This decline can be reversed by the injection of a booster dose.

## 1. Introduction

Vaccination against Covid-19 began in late December 2020 in France. By the end of 2021, the full vaccination coverage reached 77% overall ([datavaccin](https://datavaccin-covid.ameli.fr/pages/synthese/)), and 91% for people aged 18 and over. Meanwhile, the French territory was hit by three epidemic waves, defined by an acceleration of infections and hospitalizations related to severe forms of the disease, each one of them being attributed to a distinct dominant SARS-CoV-2 variant strain: the Alpha variant for the wave that peaked in March-April 2021, the Delta variant for the one that peaked in August 2021, and the Omicron variant for the one that peaked in early 2022. The latter was detected in France in early December 2021 and spread more quickly than any previous SARS-CoV-2 variant. Despite the gradual vaccine rollout, the burden of Covid-19 in France remained heavy in 2021. While vaccine coverage reached high level, part of the population remained reluctant to get the vaccine. In this context, it is essential to get reliable evidence on the protection provided by vaccines against symptomatic infections, hospitalizations and inpatient deaths related to Covid-19, on the evolution of this protection over time and after the booster dose, and on the relative effectiveness of vaccines against the different variants of concern.

At first, due to limited availability of vaccine doses, vaccines were administered in priority to healthcare workers and those most likely to develop severe forms of the disease, in particular to people aged 75 years and over, those living in retirement homes, and long-term care units. Vaccination was extended to all caregivers, and progressively to all age groups, starting with those with comorbidities, with a decreasing lower age limit for vaccine eligibility: 50 years, 18, 12 and five years (Appendix 1). Four different Covid-19 vaccine brands were used: Moderna, Pfizer/BioNTech, AstraZeneca and Janssen. A vaccination scheme was initially considered complete after two doses (or one dose for those infected by SARS-CoV-2), except for the Janssen vaccine, for which one dose was deemed sufficient. A full vaccination cycle became a sufficient condition required to get a French health pass that came into effect in June, and was first required to access events and places gathering many people. From August, the health pass was extended to grant entry into all museums, bars, restaurants, trains and other public spaces (irrespective of the number of people). The injection of a booster dose started in early September. First limited to the most fragile people and people aged 65 years or over, it became available to all professionals caring for these vulnerable people, and to all adults. This booster became mandatory for the health pass to remain valid as of December 15, 2021, and January 15, 2022, for those aged respectively 65 years or over, and 18 years or over.

Vaccination coverage progressed at unprecedented speed worldwide, providing growing evidence on vaccine effectiveness against the risk of symptomatic infections and severe forms of Covid-19 in the real world. The early estimates showed a good effectiveness of the first vaccine dose against the Alpha variant, with 61% (CI95%: 51-69) protection against symptomatic infections in people over 70 years of age, one month after the first dose of the Pfizer/BioNTech and AstraZenecca vaccines (Lopez Bernal et al., 2021a). In people aged 80 years or over, vaccinated with Pfizer/BioNTech (resp. AstraZenecca), an additional reduction in the risk of hospitalization of 43% (33-52) (resp. 37%; 3-59) was observed; thus resulting in an effectiveness against the risk of hospitalization of about 80% after a single dose of one of the two vaccines. The protection provided by vaccination against the Alpha variant, increased after the second vaccine dose, reaching 89% (85-93) protection against symptomatic infections in people 80 years or older, with the Pfizer/BioNTech vaccine (Lopez Bernal et al., 2021a). Two weeks after the second dose, vaccination provided a reduction in the risk of severe forms of Covid-19 of 92% (91; 93) for vaccination with the Pfizer/BioNTech vaccine, 96% (94-97) with the Moderna vaccine, and 96% (65-99) with the AstraZeneca vaccine, in people aged 75 years or older in France (Botton et al., 2021). However, this protection seemed lower against the Delta variant (84%; 75-90) (EPI-PHARE, 2021). Vaccine effectiveness against symptomatic cases in individuals aged 16 years was lower by 12 to 19% with the Delta variant as compared with the Alpha variant, after one vaccine dose. Yet, these differences were smaller after two doses: 88% (85-90) with the Alpha variant and 80% (77-82) with the Delta variant (Lopez Bernal et al., 2021b).

Evidence started pointing towards a significant decline over time in vaccine effectiveness against symptomatic infections, but to a lesser extent against severe cases (Thomas et al., 2021; Tartof et al., 2021; Goldberg et al., 2021). Vaccine effectiveness against symptomatic Delta variant infections dropped, 20 weeks after vaccination, to 47% (45–50) and 70% (69-71), for AstraZeneca and Pfizer/BioNTech vaccine respectively, with a stronger decline in those aged 65 years or older, but with a lower decline against hospitalizations (Andrews et al., 2021). However, studies find that the booster dose restores similar (or even better) levels of protection against symptomatic infections and severe cases than those prior to the waning of immunity (Bar-On, et al.; 2021; Andrews, et al., 2021b; Matiuzzi and Lippi, 2022).

This paper provides with complementary insights on vaccine effectiveness and its evolution over the year 2021 in the French context, using unprecedented data matching three National exhaustive databases containing exhaustive information on Covid-19 screening (SI-DEP), vaccination (VAC-SI) and hospitalizations (SI-VIC) in France. We used data from January 1^st^ 2021 to December 12, 2021, thus stopping our analysis prior to the exponential growth of the Omicron variant. We focused on people aged 50 years or over, a population that concentrates the most severe forms of the disease and that became eligible for vaccination and then booster dose at an earlier stage (Appendix 1). We performed a two-step analysis of vaccine effectiveness against severe forms of Covid-19 and estimated: *(i)* vaccine effectiveness against symptomatic forms of Covid-19; *(ii)* vaccine protection against the risk of hospitalization and death in individuals with symptomatic forms of Covid-19. In particular, we studied the evolution of vaccine protection under the combined effect of the emergence of the Delta variant, and of the decrease in immunity over time after the completion of the primary vaccination scheme. We additionally estimated the contribution of the booster dose in restoring a significant level of protection.

## 2. Material and methods

### 2.1. Study design

We used a two-step analysis to estimate vaccine effectiveness against severe forms of Covid-19, defined as leading to hospitalizations, intensive care units (ICU) admissions, or inpatient deaths. First, we used a test-negative case–control design (Jackson and Nelson, 2013) to estimate vaccine effectiveness against symptomatic Covid-19 infections. This method, already used in the Covid-19 pandemic (Lopez Bernal et al., 2021a and 2021b; Stowe et al., 2021; Tenforde et al., 2021; Chung et al., 2021), relies on the comparison of vaccination statuses between cases (individuals with confirmed SARS-CoV 2 infection) and controls (individuals who do not test positive for SARS-CoV-2 infection), with cases and controls tested after reporting symptoms suggestive of Covid-19.

Then, we performed a survival analysis among individuals with symptomatic forms of Covid-19, to evaluate a possible additional risk reduction provided by vaccination against severe forms of the disease.

### 2.2. Data sources

Three National databases created to monitor the epidemic and the vaccination campaign were matched together. This study provides the first use of this matched dataset in an international peer-reviewed journal.

SI-VIC, the information system for monitoring victims of attacks and exceptional health situations, provides, for people infected with SARS-CoV-2, the daily number of hospitalizations in general wards and ICU, and the number of inpatient deaths. The diagnosis of infection relies on RT-PCR testing or thoracic CT scanning. This reporting system, maintained by the ANS (*Agence du Numérique en Santé*), is exhaustive and covers all healthcare structures (public and private) over the French territory.

SI-DEP, the screening information system, provides the daily number of tests performed (RT-PCR, serology and antigenic tests) for SARS-CoV-2 and the results of these tests. This database, maintained by the AP-HP (*Assistance Publique - Hôpitaux de Paris*), is exhaustive for all tests performed on the French territory (but self-tests). Since mid-2020, PCR testing was available to the population without prescription and covered by national health insurance (Appendix 1). As of January 2021, a molecular screening was performed on all RT-PCR positive samples: first to identify known variant strains (wild-type, alpha, beta, gamma); then, from June 2021, to identify some key mutations (E484K, E484Q, L452R). The presence or absence of symptoms in tested individuals should be systematically reported, but this information is missing for 20% of the RT-PCR tests performed in 2021.

VAC-SI, the Covid-19 vaccine information system, maintained by the CNAM, the French national Health Insurance (*Caisse Nationale d’Assurance Maladie*), provides the number of administrated vaccines and vaccinated persons on the French territory. This dataset covers nearly the entire French population (all those affiliated to the French Health Care System [Social security]), whether vaccinated or not, and all individuals vaccinated in France. This database contains information on vaccination (dates of injection, vaccine brand name), and information on vaccine priority populations (presence of comorbidities, healthcare professionals or social workers, retirement homes residents).

To match these databases, a pseudonym (non-meaningful character string identifying each person) was generated from the concatenation and encryption of identifying information (surname, first name, sex and date of birth). The pseudonym (but not the identifying information) is present in all the databases transmitted to the Statistics office of the French Ministry for Solidarity and Health (DREES) for statistical use, which allows the matching of data on screening, hospitalization and vaccination at the individual level. However, matching imperfections may remain (DREES, 2021; Appendix 5).

The deployment of these three databases was authorized by the French Data Protection Authority (*Commission Nationale Informatique et Libertés*). No consent of the patients is required, and the patients must be informed of their right to access, modify, rectify and delete any data concerning them. The French Ministry for Health is accountable to implement legal, technical and organizational measures to guarantee data protection.

### 2.3. Study period and study population

The data used were extracted on the 11^th^ of January 2022, for observations from January 1^st^ to December 12, 2021. In the last week of the study period, the Omicron variant represented less than 10% of the positive RT-PCR tests with molecular screening. We excluded the last weeks of December when the Omicron variant started its exponential growth, as more data is needed to estimate vaccine effectiveness against this emerging variant.

We included tests performed on individuals: *(i)* aged 50 years or over, a population that concentrates the most severe forms of the disease and that became eligible for vaccination at an earlier stage; *(ii)* tested by RT-PCR, to focus on recent infections for which the causative variant could be identified; *(iii)* reporting symptoms in the seven days prior to the time of screening. When several positive tests were associated with the same pseudonym, we considered those performed less than 15 days apart as part of the same infectious episode. We thus identified distinct infectious episodes, and we included in the analysis only one positive test per episode and per individual (in priority a test with molecular screening). For a given individual, we also excluded the negative tests that had been performed within 15 days of a confirmed infectious episode.

We included only individuals present in the VAC-SI register and with non-missing information about the presence or absence of comorbidities considered for prioritizing vaccine administration in the 2021 French vaccination campaign. For our analysis, we defined four vaccination statuses: unvaccinated, one dose (D1), full primary vaccination cycle without (D2) or with booster (DB). They were further refined according to the time since the injection and the vaccine brand. In particular, individuals vaccinated with the Janssen vaccine completed their primary vaccination scheme after only one dose. The vaccination status was estimated as of the date of RT-PCR screening. In France, a full primary vaccination status is also achieved after only one dose in case of a confirmed past infection in the three months preceding the first dose or in the month following that dose (set aside the Janssen vaccine). We excluded all individuals with a confirmed SARS-CoV-2 infection at least 60 days prior to the time of screening (in coherence with the [European Surveillance System definition](https://www.ecdc.europa.eu/en/publications-data/reinfection-sars-cov-2-implementation-surveillance-case-definition-within-eueea) of suspected cases of SARS-CoV-2 reinfection), in order to estimate vaccine effectiveness in fully susceptible individuals. This choice implies that a full primary vaccination cycle only relates to the injection of two doses. We excluded individuals with atypical vaccination schemes: those vaccinated with two doses from two different vaccines when one of them was the Janssen vaccine, and those who never received their second injection of a bi-dose vaccine (they were excluded 28 days after their first dose).

To study severe forms of Covid-19, we included the inpatients aged 50 years or over that were present in all three databases (SI-VIC, SI-DEP and VAC-SI). We applied the filters already listed above on data from vaccination and screening. In addition, we excluded inpatients: *(i)* infected by SARS-CoV-2 but whose admission was not attributable to Covid-19 (persons hospitalized for other conditions may have been systematically screened); and *(ii)* with no symptomatic positive RT-PCR test recorded within 15 days before hospital admission, or within two days after. In case of symptomatic positive RT-PCR test within two days after admission, inpatients are reclassified as tested on the day of admission to avoid negative durations in the survival analysis, and kept in the analysis.

### 2.4. Statistical analysis

We used a test-negative case-control design to estimate vaccine effectiveness against symptomatic Covid-19. Symptomatic positive individuals (respectively positive to a given variant for the analysis by variant) were randomly matched to controls (symptomatic negative individuals) on age (ten-year age brackets), sex, area of residence (NUTS-3 level), week of testing and presence or absence of a comorbidity qualifying for prioritization in the vaccination campaign. The odds ratios (OR) were estimated using a conditional logistic regression, and vaccine effectiveness (*VE*(*S*+)) was given by the following formula: *VE*(*S*+) = 1 − *OR* (Bond et al., 2017).

We then estimated the risk of severe outcome (hospitalization, ICU admission, or death) among individuals with RT-PCR-confirmed SARS-CoV-2 symptomatic infection, according to their vaccination status. We fitted a Cox survival model on the time interval between the date of the test and the date of the hospital admission associated with the severe outcome, if any, or the end of the follow-up period. The latter was censored at 15 days post-test or at the end of the study period (December 12, 2021). A hazard ratio of hospitalization (respectively ICU admission or death) was then estimated according to the vaccination statuses (*HR*(*H*|*S* +)), controlling for the same variables as those used in the case-control study.

This two-step analysis allowed calculating the vaccine effectiveness against severe forms of Covid-19, *VE*(*H*), *VE*(*ICU*), *VE*(*D*), for hospitalization, ICU admission and inpatient death respectively, considering that, in addition to the risk reduction against symptomatic forms, vaccination also provides a risk reduction in the development of severe disease in symptomatic individuals. Vaccine effectiveness against severe forms of Covid-19 was deduced from the following formula: *VE*(*i*) = 1 − *OR*(*S*+) *\* HR*(*i*|*S*+), where *i* refers to either hospitalization, ICU admission or inpatient death.

We estimated the evolution of vaccine effectiveness over time, according to the time elapsed since each vaccine dose. We provided estimates up to six months after the second dose, except for the variant-specific analysis, that we truncated three months after the second dose. Indeed, for all variants but the Delta one, the variant-specific incidence was very low when the population reached this duration after a full vaccination scheme.

## 3. Results

### 3.1. Description of the study population

Over the period from January 1^st^ to December 12, 2021, 8 881 107 individuals were tested by RT-PCR and reported symptoms at the time of screening. 2 413 356 (27%) of them were aged 50 years or over and 2 024 773 (84%) of the latter were successfully linked to vaccination data with non-missing data on comorbidities. We excluded 44 604 individuals with unusual vaccination schemes, and 67 134 individuals with a known past infection prior to the time of screening. Of these remaining individuals, 437 694 (23%) were tested positive for SARS-CoV-2.

The study population for the test-negative design analysis consisted of 432 117 positive cases (5 577 cases were excluded because of the lack of similar controls) and 864 234 controls (two matched controls for each case). Almost none of them was vaccinated at the beginning of the year 2021, 50% had received one dose by May, the 20^th^ of 2021, and two doses by July, the 2^nd^ of 2021. By the end of 2021, 86% of them had completed their primary vaccination cycle and 46% of them had received a booster dose. The distribution of vaccination statuses over time differed between cases and controls, with an earlier start of the primary vaccination scheme in the latter (Figure 1, Appendix 2).

**Figure 1.**
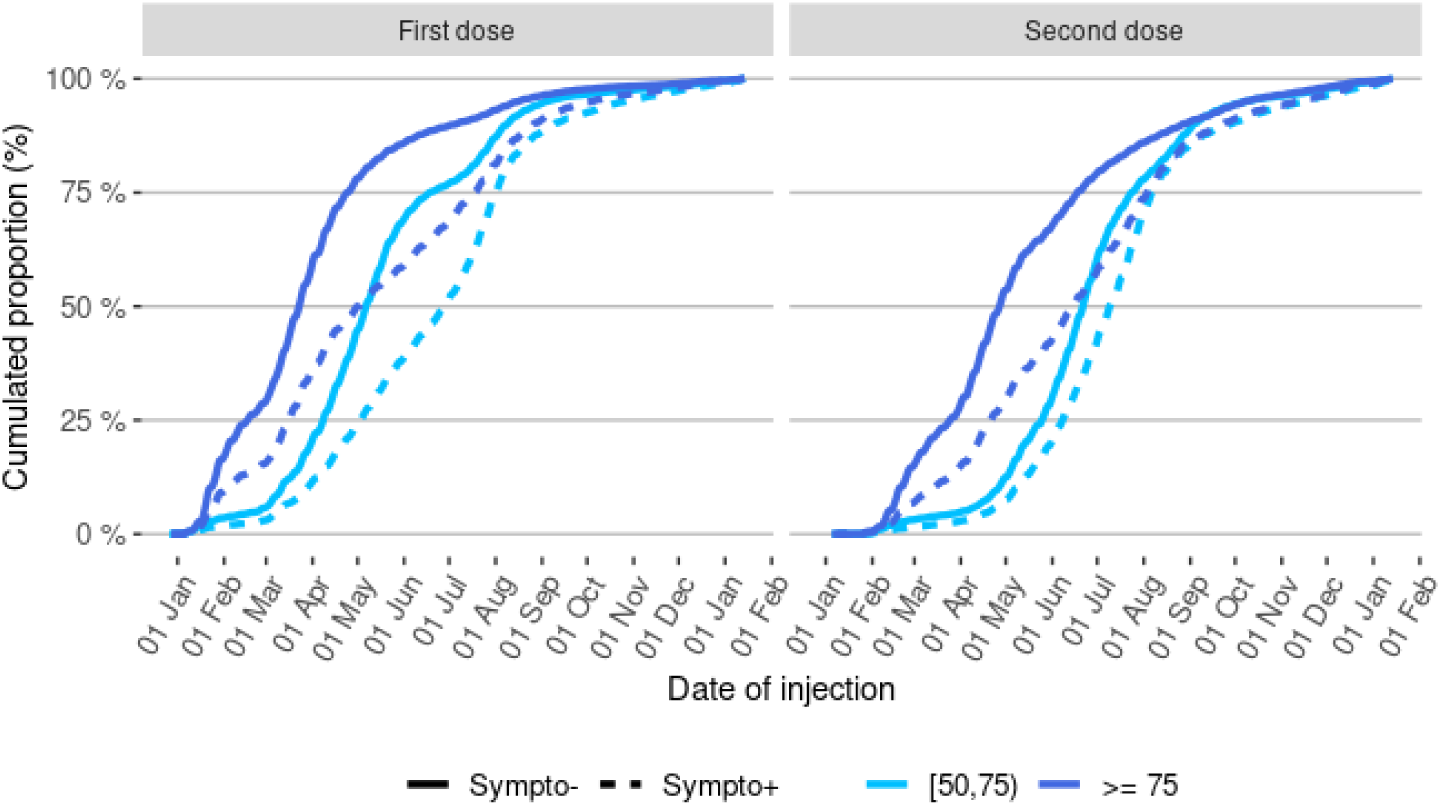
Distribution of injection dates for the first and second vaccine doses in control and cases, by age-group Abbreviations: Sympto+ (cases): symptomatic individuals with a laboratory confirmed SARS-CoV-2 infection (cases). Sympto- (controls): individuals with symptoms non-related to SARS-CoV-2 infection.

Among the symptomatic positive cases in the sample, 67% had not yet been vaccinated when they got tested, 15% were older than 75 years, 52% were women, and 63% had a comorbidity (Appendix 2).

The study population for the survival analyses consisted of the 437 694 persons with confirmed SARS-CoV-2 symptomatic infection, among which there were 44 615 hospitalizations, 12 050 ICU admissions and 7 476 inpatient deaths recorded in SI-VIC. We did not consider the 4 813 hospitalizations that did not meet the criteria listed in the study population section.

### 3.2. Vaccine effectiveness of the primary vaccination cycle

Among those aged 50 years and over, vaccine effectiveness against symptomatic infection grew quickly even after only one dose and further increased after the completion of the primary vaccination cycle. It reached 26% (24-28) two weeks after the first dose and 45% (43 – 47) one month after, and it peaked at 82% (81 – 83) two weeks after the second dose (Figure 2). However, in the early days after the first dose, before protective immunity has been reached, we observe an increased risk of symptomatic infection in vaccinated persons versus comparable unvaccinated persons.

**Figure 2.**
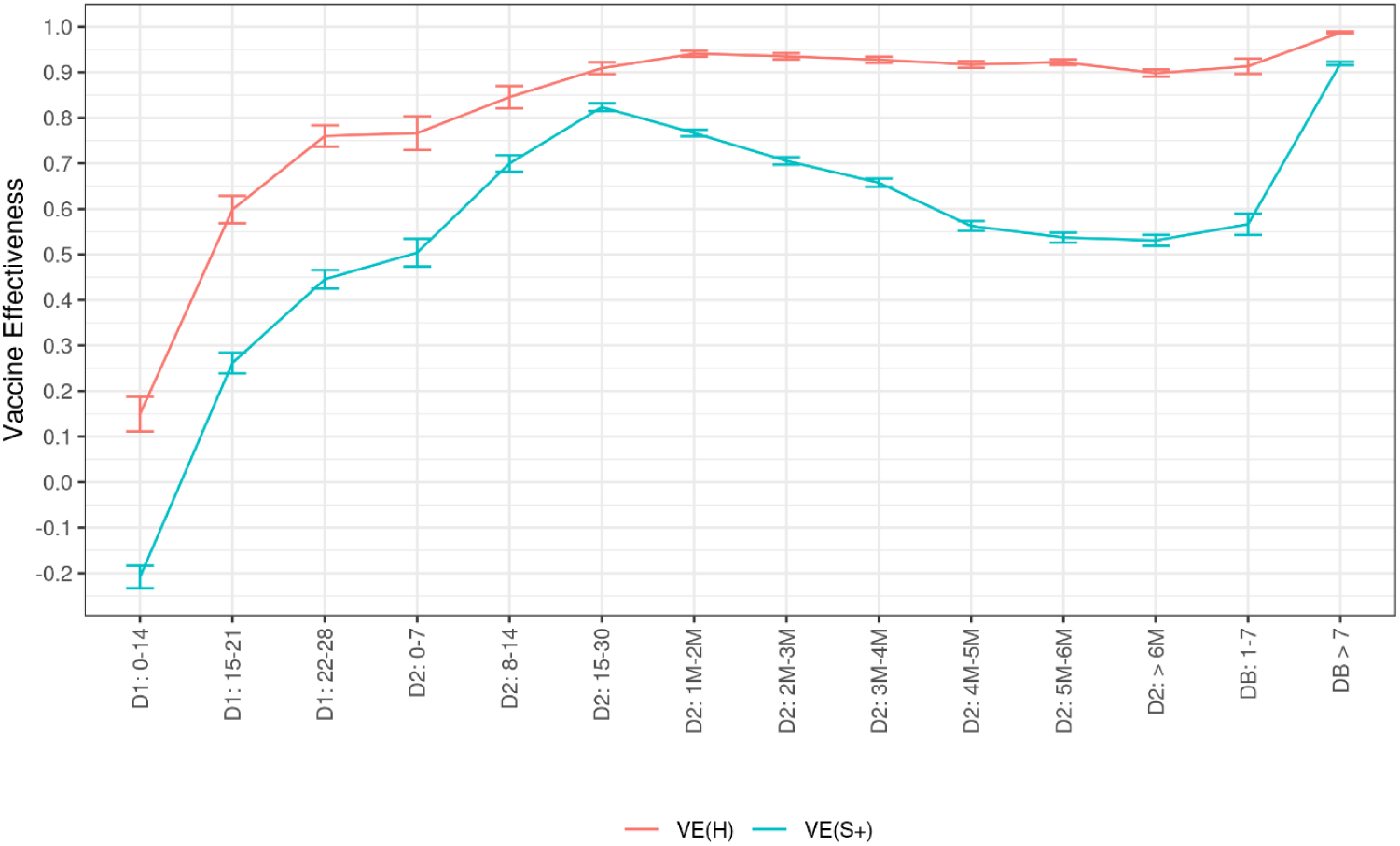
Covid-19 vaccine effectiveness against symptomatic infections and hospitalizations among persons aged 50 years or over, according to the time elapsed since the injection of each vaccine dose, data collected from January 1^st^ to December 12, 2021 Abbreviations: D1: first vaccine dose. D2: second vaccine dose. DB: booster dose. M: month. S+: symptomatic infection. H: hospitalization. VE: vaccine effectiveness. The numbers in the x-axis indicate the time (in days or months) elapsed since the injection of the dose of interest.

The risk of hospitalization, ICU admission, and inpatient death of infected symptomatic individuals decreased as the vaccination cycle progressed (Figure 3). Among them, vaccination provided more than 75% (72-77) risk reduction against hospitalizations and ICU admissions, and 54% (44-63) risk reduction against inpatient deaths, one month after the injection of the second dose.

**Figure 3.**
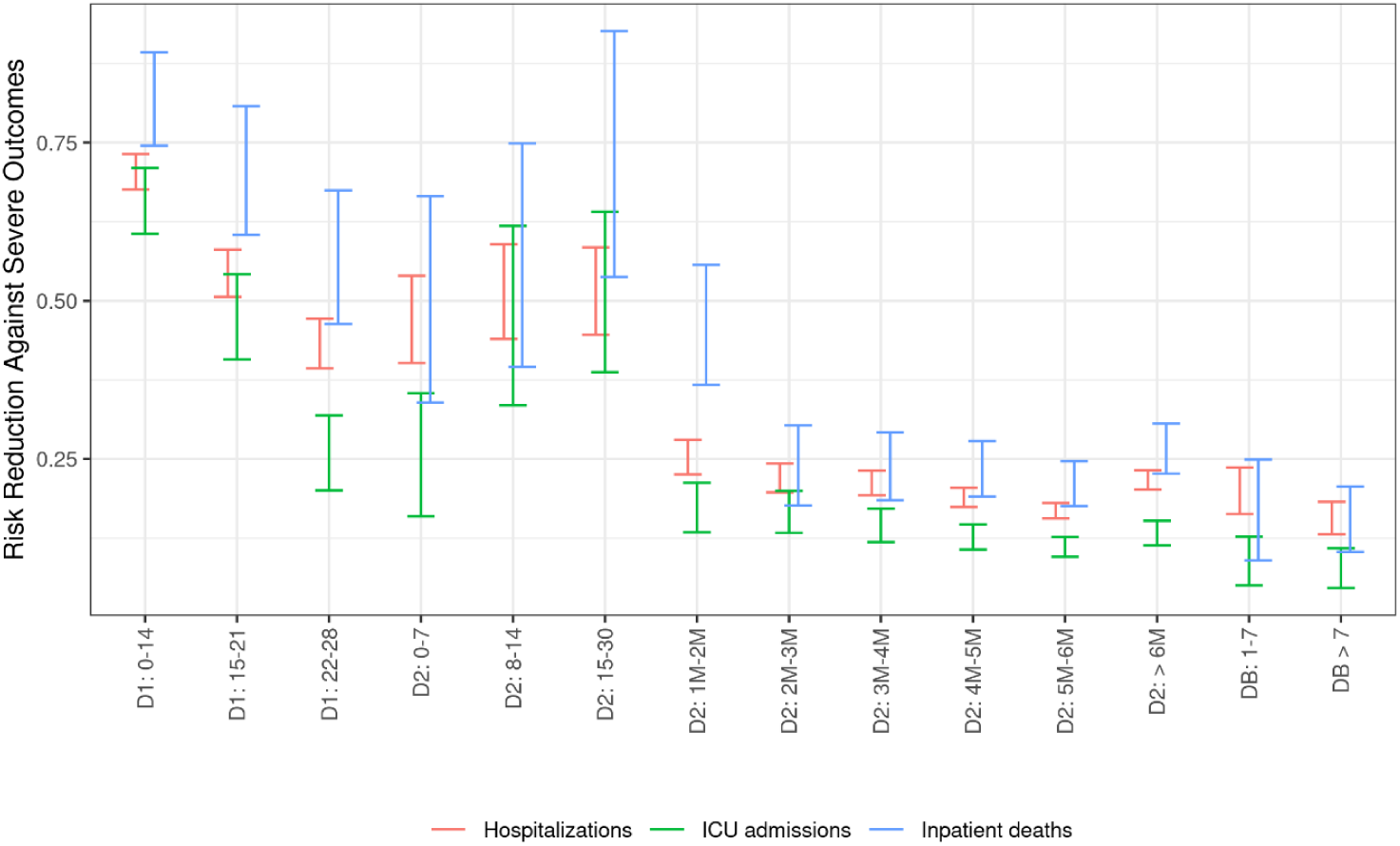
Risk reduction against Covid-19 severe outcomes (hospitalizations, ICU admissions and inpatient deaths) among persons aged 50 years or over, according to the time elapsed since the injection of each vaccine dose, data collected from January 1st to December 12, 2021 Abbreviations: D1: first vaccine dose. D2: second vaccine dose. DB: booster dose. M: month. a The numbers in the x-axis indicate the time (in days or months) elapsed since the injection of the dose of interest.

**Figure 4.**
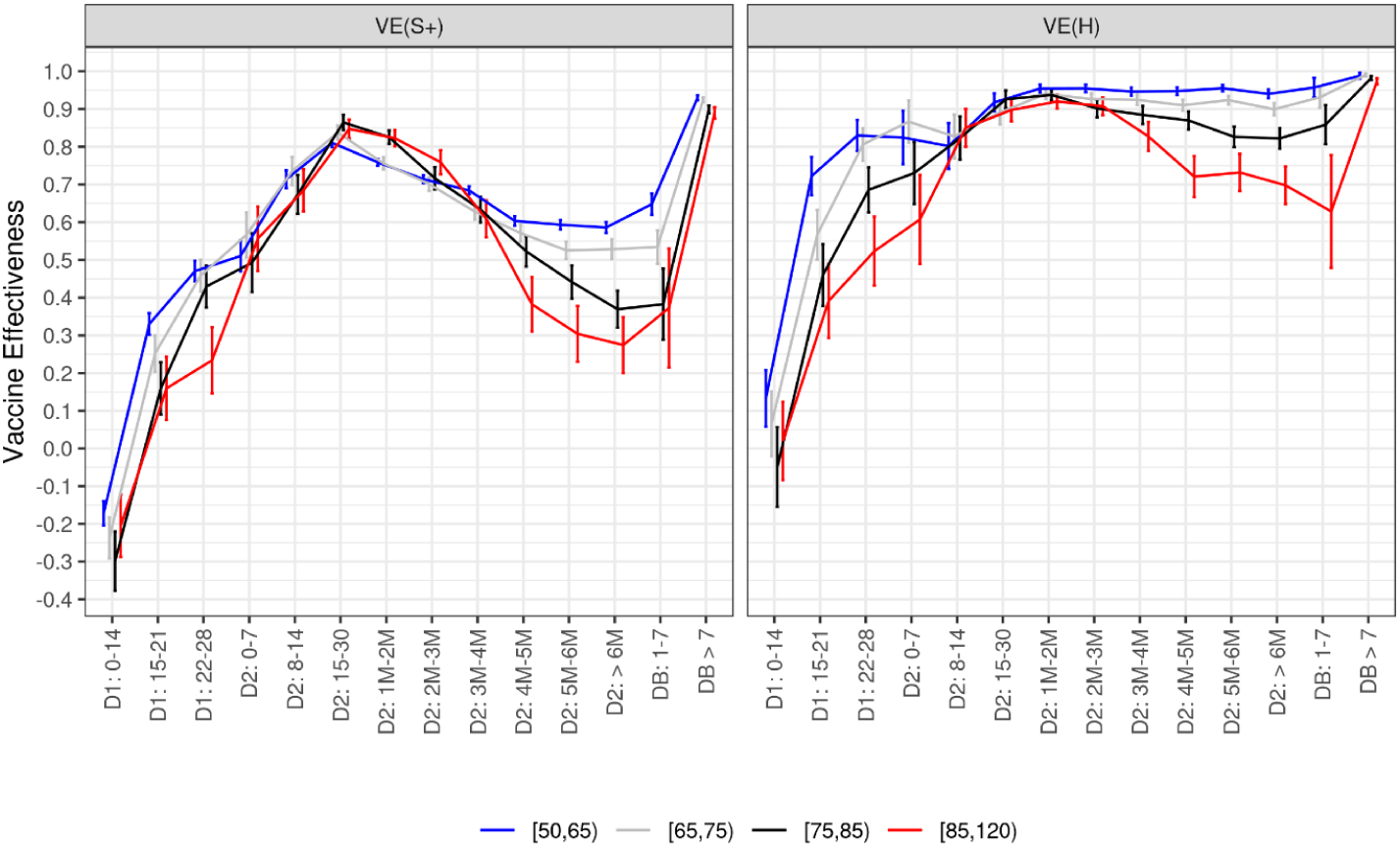
Covid-19 vaccine effectiveness against symptomatic infections and hospitalizations by age, according to the time elapsed since the injection of each vaccine dose, data collected from January 1^st^ to December 12, 2021 Abbreviations: D1: first vaccine dose. D2: second vaccine dose. DB: booster dose. M: month. S+: symptomatic infection. H: hospitalization. VE: vaccine effectiveness. The numbers in the x-axis indicate the time (in days or months) elapsed since the injection of the dose of interest.

We then combined the estimates of vaccine effectiveness against symptomatic infections with the additional protection provided by vaccination against severe forms of the disease in those experiencing symptomatic infections. We thus obtained a vaccine effectiveness that peaked at 94% (93 – 95) against hospitalizations, 96% (95 – 97) against ICU admissions and 89% (87 – 91) against inpatient deaths after a primary vaccination cycle (Figure 2).

### 3.3. Decline in vaccination effectiveness over time, before the booster shot

Among persons aged 50 years or over, the vaccine effectiveness against symptomatic infections peaked in the first month after the second dose, before declining sharply (Figure 2), and falling to 53% (52–54) within six months. However, the additional risk reduction for ICU admissions and inpatient deaths among symptomatic individuals decreased only very little over time (Figure 3), and remained constant for hospitalizations. As a result, vaccine effectiveness against severe disease declined less and slower. It was still about 90% (89–91) against the risk of hospitalization, more than six months after the second injection.

The booster dose seemed very efficient in restoring vaccine effectiveness to levels even higher than ever: reaching 92% against symptomatic forms and 99% against hospitalizations.

### 3.4. Vaccination effectiveness by age and comorbidities

Vaccine effectiveness against symptomatic infections was similar at the beginning of the vaccination cycle and up to five months after two doses for people aged 50 years or over with comorbidities, but was six percentage points lower after six months (Appendix 3). When studying severe Covid-19 outcomes, the presence of comorbidity, independently of vaccination status, was associated with a higher risk of complications with a hazard ratio of 1·57 (1·54-1·61) for hospitalization, 1·72 (1·65-1·79) for ICU admission, and 1·35 (1·27-1·43) with inpatient death. However, the vaccine reduced the risk of severe outcome in a similar way for people with comorbidities than for all people aged 50 years or more. As a result, vaccine effectiveness against severe Covid-19 after a full vaccination scheme appeared very similar for people with comorbidities. The booster dose then restored similar and high protection levels in both categories.

Vaccine effectiveness against symptomatic infections and severe cases showed no significant differences between age groups (for people aged 50 years or over) right after the completion of the primary vaccination cycle. However, vaccine effectiveness against symptomatic cases, which significantly declined for all age groups, decreased stronger among the elderly. After six months, it dropped to 59% (57-60) in people aged 50 to 74, to 37% (32-41) in people aged 75 to 84, and to 27% (20-34) in those aged 85 and over. Vaccine effectiveness against hospitalizations declined moderately for all age groups but those aged 85 years or over, for whom it dropped to 70% (64-75) after six months. Again, high vaccine effectiveness in these frail and older populations was restored after the booster dose, up to levels comparable to those obtained in the younger age groups (97%; 96-98).

### 3.5. Vaccination effectiveness against different variants

Vaccine effectiveness did not differ significantly against the Alpha variant and the wild type SARS-CovCov-2, reaching 91% (90-92) and 92% (88 – 96) respectively, 15 days after the second injection (Figure 5 and Table 4 in Appendix 3). In comparison, vaccine effectiveness against the Beta/Gamma and Delta variants were lower against symptomatic infections, at all stages of the vaccination course, peaking respectively at 84% (78–90) and 79% (77–80), 15 days after the second injection. Point estimates suggest a lower effectiveness against the Beta/Gamma variants than against the Delta one after one month, but these differences are not significant. A decrease in vaccine effectiveness against symptomatic infections is observed over time for all variants (but the wild type). In contrast, vaccine effectiveness against hospitalization does not appear to decrease in the first three months, regardless of the variant. At the peak and from two to three months after the second dose, this effectiveness is significantly higher against the Alpha variant than against the Delta one by four percentage points.

**Figure 5.**
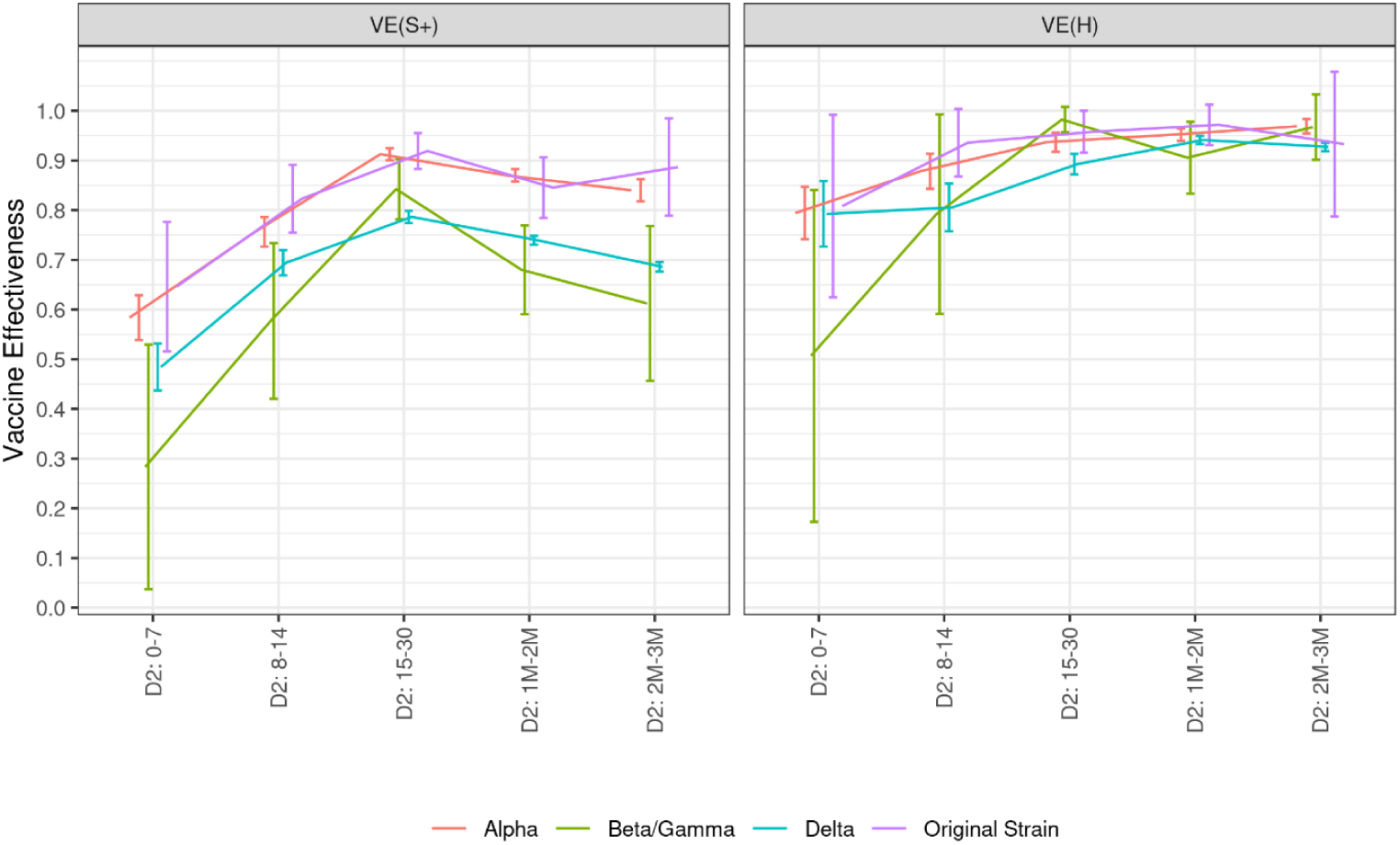
Covid-19 vaccine effectiveness against symptomatic infections and hospitalizations related to various variants of concern, according to the time elapsed since the injection of each vaccine dose, data collected from January 1^st^ to December 12, 2021 Abbreviations: D1: first vaccine dose. D2: second vaccine dose. DB: booster dose. M: month. S+: symptomatic infection. H: hospitalization. VE: vaccine effectiveness. The numbers in the x-axis indicate the time (in days or months) elapsed since the injection of the dose of interest.

## 4. Discussion

We have used three large and exhaustive datasets on Covid-19 screening, vaccination and hospitalizations in France to assess vaccine effectiveness over the year 2021. On the study population aged 50 years or more, we found a high vaccine effectiveness against symptomatic and severe forms of the disease, which increased as one progressed through the vaccination scheme. At its peak, the vaccine effectiveness of a primary vaccination cycle (without booster) reached 85% against symptomatic forms and 90% against severe forms, with little difference between age groups, and no difference for individuals with comorbidities.

Vaccine effectiveness against symptomatic forms then decreased over time after the completion of the primary vaccination cycle, dropping to 57% in those aged 50 years or over six months after completion of the vaccination cycle and to 39% in those aged 85 years or over. However, protection against severe forms declined much slower and vaccine effectiveness remained high (84% six months after completion of the vaccination course), except in the oldest age group where protection over time dropped stronger (70% after six months). These estimates are consistent with observational studies in other countries.

When the time elapsed since vaccination reached several months in the first vaccinated individuals, the Delta variant had become predominant. Our findings show that the drop in vaccine protection over time seems mainly due to a decline in vaccine protection, rather than to a greater capacity of the Delta variant to escape vaccine protection. Indeed, we found that vaccine effectiveness was only slightly lower against the Delta variant than against the wild type and Alpha variant. In addition, in the first three months after vaccination, a similar drop in vaccine effectiveness was observed for all variants, which suggests that immunity waning is not specific to the Delta variant. If the reduction in vaccine protection against the variants under study remains limited, our analyses were conducted prior to the rapid spread of the Omicron variant in France, for which vaccine escape may be of greater concern. Estimates on that matter, though still little documented to date, point towards very little (or no) protection against symptomatic infections (Buchan et al. 2022; Andrews et al. 2021c), but partially restored protection with a booster (Willet et al. 2022) and a limited protection against severe cases (Collie et al. 2021).

We showed that the highest levels of protection against both symptomatic and severe forms of the disease were obtained one week after receiving a booster dose, without distinction on age groups. This confirms that the booster dose is essential to restore a high level of protection.

These real-world estimates are also consistent with laboratory analyses: although the amount of antibodies in vaccinated individuals decreases over time (Levin et al., 2021), memory B cells seem to remain numerous and capable of a better response (Goel et al., 2021), thus likely preventing severe cases. In addition, a rapid serological response was observed after the booster dose (Pfizer/BioNTech), with significantly higher antibody titers than those observed after the second dose (Ireland et al., 2022).

The exhaustiveness of the databases and the possibility to match them together to get information at an individual scale are the great strengths of this study. We thus provide evidence of vaccine effectiveness in real-world conditions over a long period. The large sample size allows us: *(i)* to provide reliable estimates of vaccine protection against severe Covid-19, which remain rare events; and *(ii)* to detail these estimates for several sub-populations and with a precise time elapsed since the injection of all vaccine doses, up to the booster dose. However, the observational nature of the data itself also brings about some limitations. We used a test-negative design in order to reduce selection biases that are difficult to measure such as health-seeking behaviour, access to testing and case ascertainment. This method has been proven useful in our study context. In particular, it allowed limiting the effect of the evolution of screening policies on the propensity of getting tested (Appendix 1). Yet, test-negative designs rely on strong assumptions, the applicability of which is difficult to assess and may have varied over the study period (Jackson and Nelson, 2013; Dean et al, 2021). The control variables that are used to limit the selection bias in the use of vaccination may not be sufficient, as a given vaccination status at a given time may reflect unobserved factors. For example, among people aged 50 years or over, those vaccinated at the beginning of the study period are likely to be more vulnerable (persons in retirement homes or with comorbidities), whereas those unvaccinated at the end of the study period are likely to be special (persons for whom vaccination is contraindicated, or persons against vaccination). In our study, we observed an increased risk of infection in the first days after vaccination, before protective immunity has been reached. This finding, observed elsewhere (Lopez Bernal et al. 2021a; Chung et al. 2021), seems to be due to a higher baseline risk of infection among those who were initially prioritized to receive the vaccine, suggesting missing unobserved controls (Appendix 4). In addition, being vaccinated could affect: *(i)* the perception of the need to be screened in case of symptoms and *(ii)* the probability of exposure to the virus, if being vaccinated leads to an increase in social interactions or to a lesser application of barrier gestures.

Beyond the biases inherent to observational studies, some limitations are linked to the data and variables available. Even though we excluded individuals with a known past infection based on virological and serological tests performed prior to the first vaccination dose, some past infections remained undetected and the vaccine effectiveness of the first dose of vaccine may partially reflect the stimulation of pre-existing immunity. The matching between the three databases was not perfect, which led to sample restrictions that could affect the representativeness of the results obtained. However, the corrections made to improve the matching between databases have not resulted in significant revisions of the results. Our findings only relate to vaccine effectiveness against symptomatic infections, but these symptoms were self-reported, without medical advice. We did not attempt to distinguish results by type of vaccine, although the durability of vaccine protection and the ability to protect against different variants may differ (EPI-PHARE, 2021; Andrews et al., 2021).

Overall, we found high levels of vaccine effectiveness against symptomatic infections and severe diseases after a full primary vaccination cycle. The protection was high against all the variants that circulated in France prior to December 2021, including the Delta variant that did not show a strong capacity to escape vaccine immunity. The decline of effectiveness overtime -which is strong against symptomatic infections but remains limited against severe diseases-, is efficiently restored by a booster dose. Our findings underscore the importance of monitoring vaccine effectiveness over time, and of maximizing the vaccine uptake of the booster dose.

## Data Availability

Data was obtained to carry out the legal missions of the DREES (health surveillance and alert, healthcare performance assessment), and therefore benefited from the legal prerogatives vested in the French Ministry for Solidarity and Health to carry out these missions.

## Acknowledgements

Frédéric Tallet^1^, Charlotte Geay^1^, Mathilde Gaini^1^, Gladys Baudet^1^, Catherine Pollak^1^, Benoit Ourliac^1^, Daniel Lévy-Bruhl^2^, Juliette Paireau^2^

1. DREES, Statistics office of the French Ministry for Solidarity and Health, Paris, France.

2. Santé Publique France, French National Public Health Agency, Saint-Maurice, France

## Author contributions

All authors conceived the study, developed the methods, performed analyses, and co-wrote the paper.

## Ethical considerations

The study was based on the analysis of pseudonymised data collected from health professionals. According to French law, such studies are not required to receive ethics committee approval, as the study was carried out as a contribution to the legal missions of the DREES (health surveillance and alert, healthcare performance assessment), and therefore benefited from the legal prerogatives vested in the French Ministry for Solidarity and Health to carry out these missions. The DREES is allowed to process personal health data in order to compute statistics, under article 65 of the law “Data processing and Liberties” (*Informatique et Libertés*) of January 6th, 1978. In that case, these statistics aim to prevent, manage and assess the consequences of the covid epidemic.

## Appendix. Supplementary materials

## Appendix 1 Milestones of the French sanitary crisis and its management

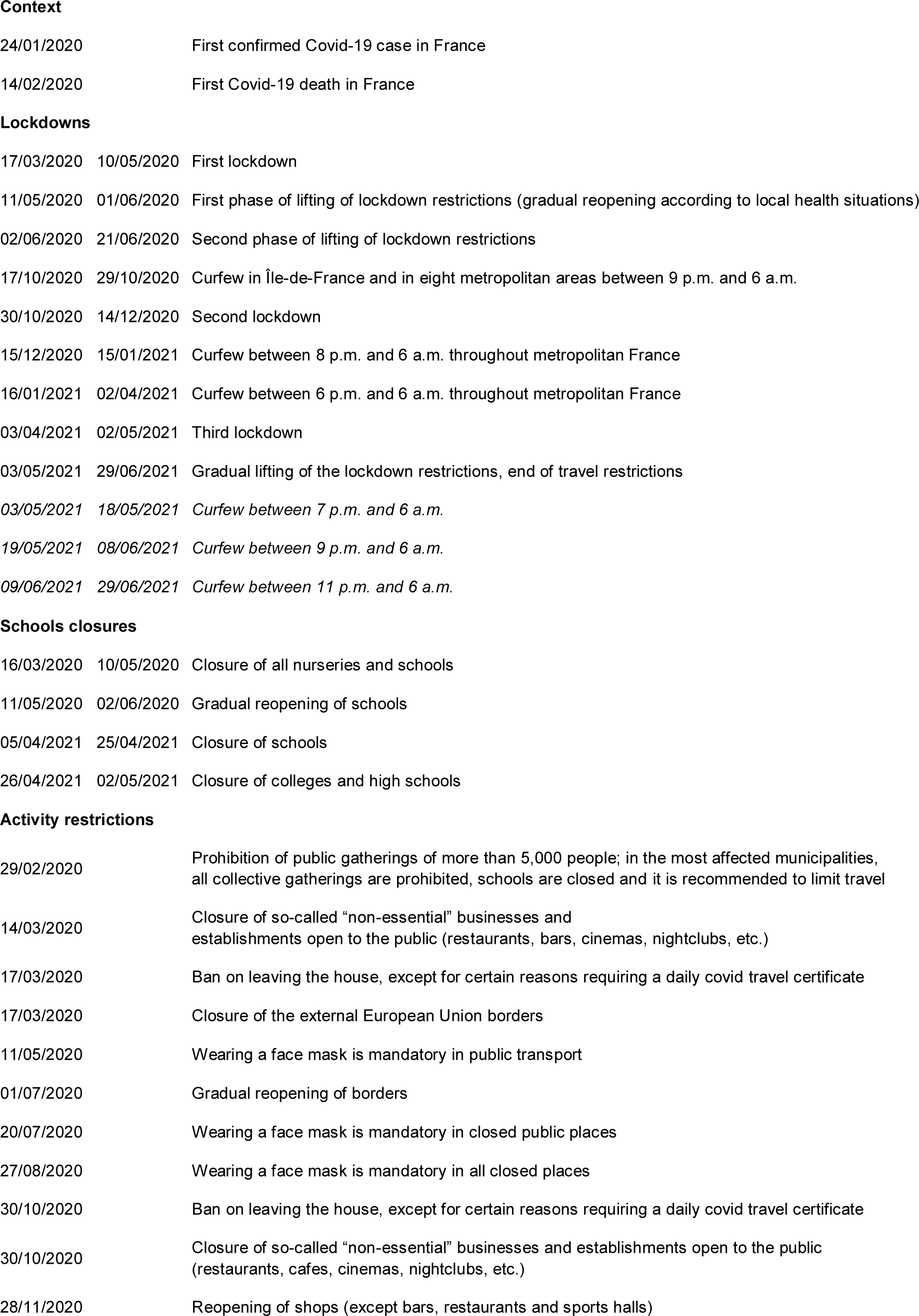

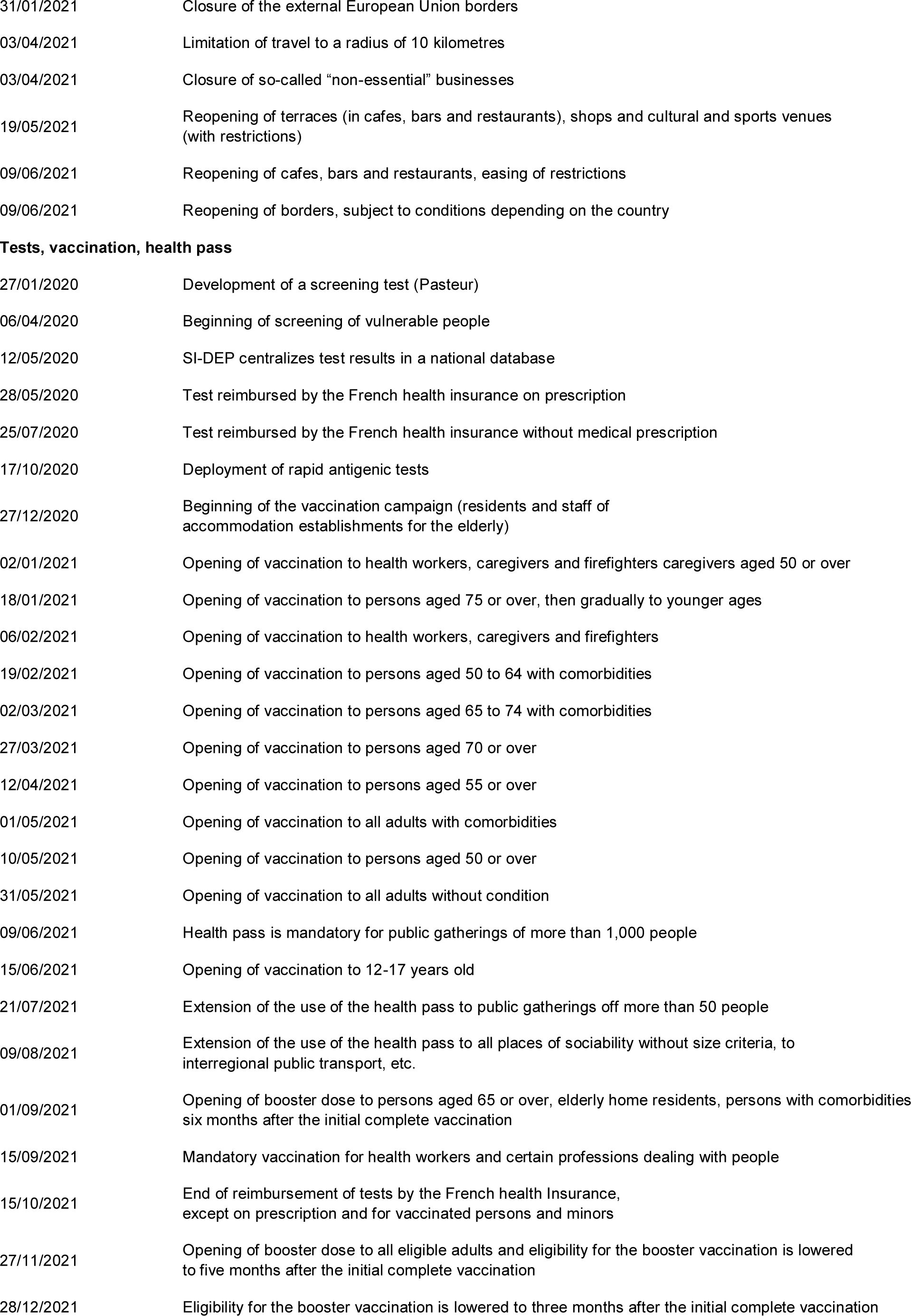

## Appendix 2 Description of the study population

**Table 1.** • Descriptive characteristics of the sample, data collected from January 1^st^ to December 12, 2021.

|  | Symptomatic negatives | Symptomatic positives | Hospitalized |
| --- | --- | --- | --- |
| Number of persons | N=1,475,037 | N=437,693 | N=45,802 |
| **Characteristic** |  |  |  |
| **Sex — no. (%)** |  |  |  |
| Female | 848,818 (57.5) | 226,306 (51.7) | 18,592 (40.6) |
| Male | 626,208 (42.5) | 211,387 (48.3) | 27,210 (59.4) |
| Missing data | 11 (<0.1) | 1 (<0.1) | 0 |
| **Age — no. (%)** |  |  |  |
| 50-64 yr | 815,682 (55.3) | 276,597 (63.2) | 14,323 (31.3) |
| 65-74 yr | 348,718 (23.6) | 95,273 (21.8) | 11,733 (25.6) |
| 75-84 yr | 174,709 (11.8) | 39,127 (8.9) | 9,690 (21.2) |
| ≥ 85 yr | 135,928 (9.2) | 26,697 (6.1) | 10,056 (22.0) |
| **Comorbidity — no. (%)** |  |  |  |
| No | 289,793 (19.6) | 162,870 (37.2) | 12,051 (26.3) |
| Yes | 1,185,244 (80.4) | 274,824 (62.8) | 33,751 (73.7) |
| **One dose — no. (%)** |  |  |  |
| No | 812,653 (55.1) | 291,036 (66.5) | 36,345 (79.4) |
| Yes | 662,384 (44.9) | 146,658 (33.5) | 9457 (20.6) |
| **One dose type — no. (%)** |  |  |  |
| Comirnaty (Pfizer/BioNTech) | 471,622 (71.2) | 100,159 (68.3) | 6,597 (69.8) |
| Spikevax (Moderna) | 55,554 (8.4) | 8,314 (5.7) | 605 (6.4) |
| Vaxzevria (AstraZeneca) | 121,270 (18.3) | 32,864 (22.4) | 1,865 (19.7) |
| Janssen | 13,938 (2.1) | 5,321 (3.6) | 390 (4.1) |
| **Two doses — no. (%)** |  |  |  |
| No | 970,735 (65.8) | 330,638 (75.5) | 41,262 (90.1) |
| Yes | 504,302 (34.2) | 107,056 (24.5) | 4,540 (9.9) |
| **Two doses type — no. (%)** |  |  |  |
| Comirnaty (Pfizer/BioNTech) | 390,933 (77.5) | 78,262 (73.1) | 3,373 (74.3) |
| Spikevax (Moderna) | 47,118 (9.3) | 6,116 (5.7) | 278 (6.1) |
| Vaxzevria (AstraZeneca) | 66,251 (13.1) | 22,678 (21.2) | 889 (19.6) |

**Figure 6.**
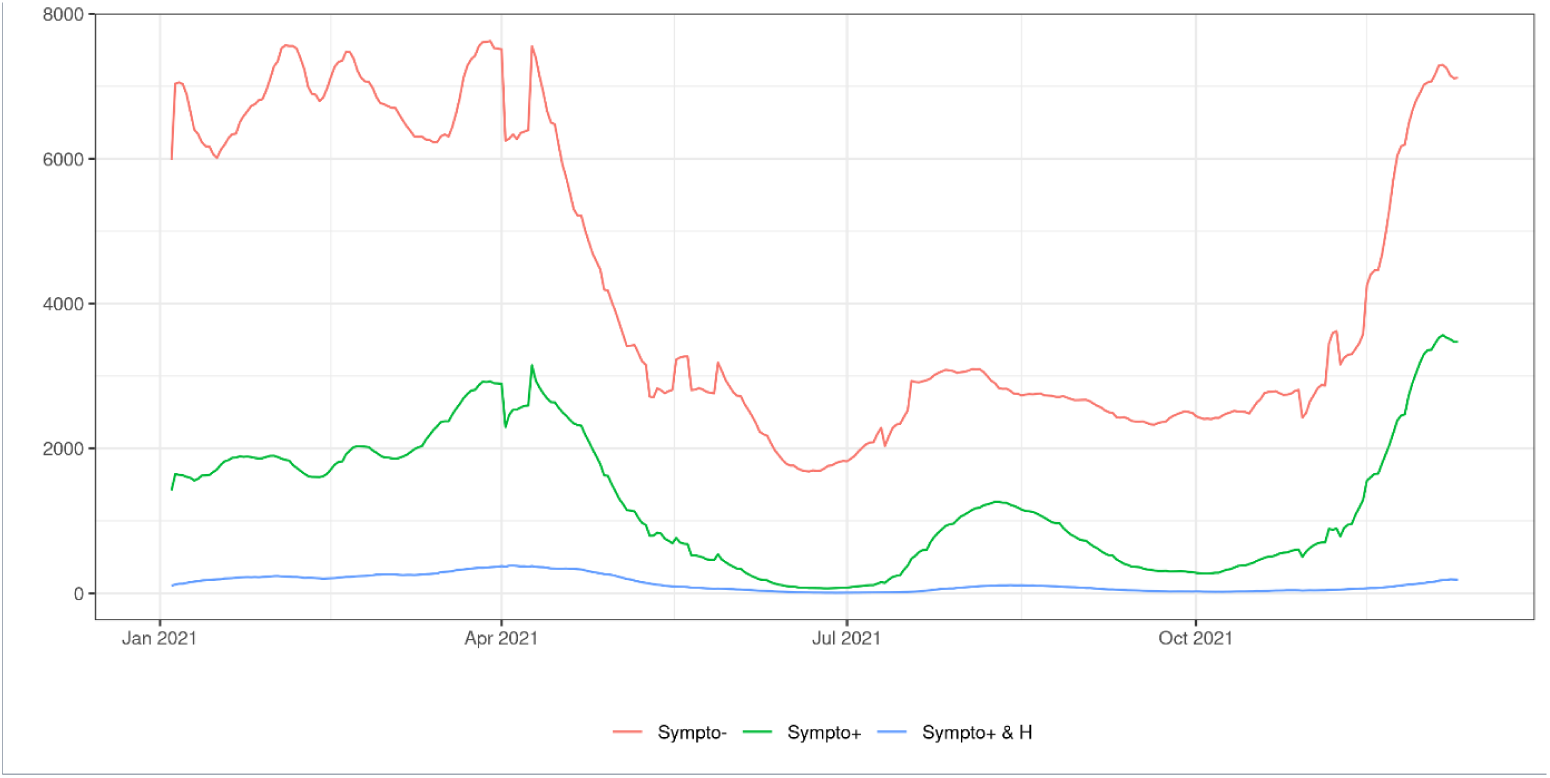
Daily counts of controls, symptomatic and hospitalized cases (averaged over the last 7 days), data collected from January 1^st^ to December 12, 2021 Abbreviations: Sympto+ (cases): symptomatic individuals with a laboratory confirmed SARS-CoV-2 infection (cases); Sympto+ & H (hospitalized cases): symptomatic individuals with a laboratory confirmed SARS-CoV-2 infection and hospital admission for Covid-19. Sympto- (controls): individuals with symptoms non-related to SARS-CoV-2 infection.

## Appendix 3

**Table 2.** Covid-19 vaccine effectiveness (in %) against symptomatic infections and hospitalizations among persons aged 50 years or over, according to the time elapsed since the injection of each vaccine dose, data collected from January 1^st^ to December 12, 2021.

| Age | 50 years or over |  | 50 years or over with comorbity |  |
| --- | --- | --- | --- | --- |
| Vaccination status | VE(S+) | VE(H+) | VE(S+) | VE(H+) |
| D1: 0-14 | -20.8 (-23.3--18.3) | 15.0 (11.1-18.8) | -5.5 (-8.5--2.4) | 28.0 (24.2-31.7) |
| D1: 15-21 | 26.2 (23.9-28.4) | 59.9 (56.8-62.9) | 28.5 (25.5-31.6) | 61.6 (58.4-64.9) |
| D1: 22-28 | 44.5 (42.5-46.6) | 76.0 (73.7-78.3) | 44.6 (41.9-47.3) | 76.3 (73.8-78.9) |
| D2: 0-7 | 50.4 (47.4-53.4) | 76.7 (73.0-80.4) | 57.6 (54.0-61.2) | 80.0 (76.5-83.5) |
| D2: 8-14 | 70.0 (68.1-71.8) | 84.5 (82.1-87.0) | 71.0 (68.6-73.5) | 85.6 (83.0-88.2) |
| D2: 15-30 | 82.4 (81.5-83.2) | 90.9 (89.6-92.2) | 82.9 (81.7-84.1) | 91.0 (89.5-92.4) |
| D2: 1M-2M | 76.7 (76.0-77.4) | 94.1 (93.4-94.8) | 78.4 (77.4-79.3) | 94.5 (93.9-95.2) |
| D2: 2M-3M | 70.5 (69.7-71.4) | 93.5 (92.8-94.2) | 69.9 (68.5-71.2) | 93.2 (92.4-94.0) |
| D2: 3M-4M | 65.7 (64.8-66.7) | 92.7 (92.0-93.4) | 66.0 (64.6-67.4) | 92.5 (91.6-93.3) |
| D2: 4M-5M | 56.2 (55.2-57.3) | 91.7 (91.0-92.4) | 56.2 (54.5-57.9) | 91.4 (90.6-92.2) |
| D2: 5M-6M | 53.7 (52.6-54.8) | 92.2 (91.6-92.8) | 49.6 (47.8-51.5) | 91.2 (90.4-91.9) |
| D2: > 6M | 53.1 (51.9-54.3) | 89.8 (89.1-90.6) | 47.0 (44.9-49.0) | 88.1 (87.0-89.1) |
| DR: 1-7 | 56.6 (54.3-59.0) | 91.3 (89.7-93.0) | 51.7 (47.9-55.5) | 90.2 (88.1-92.2) |
| DR > 7 | 91.9 (91.6-92.3) | 98.7 (98.5-99.0) | 90.5 (89.9-91.1) | 98.4 (98.2-98.7) |

**Table 3.** Covid-19 vaccine effectiveness (in %) by age group against symptomatic infections and hospitalizations among persons aged 50 years or over, according to the time elapsed since the injection of each vaccine dose, data collected from January 1^st^ to December 12, 2021.

| Age | 50-64 yr |  | 65-74 yr |  | 75-85 yr |  | ≥ 85 yr |  |
| --- | --- | --- | --- | --- | --- | --- | --- | --- |
| Vaccination status | VE(S+) | VE(H+) | VE(S+) | VE(H+) | VE(S+) | VE(H+) | VE(S+) | VE(H+) |
| D1: 0-14 | -17.2 (-20.4--14.0) | 13.3 (5.8-20.8) | -23.7 (-29.2--18.2) | 6.5 (-2.1-15.2) | -29.9 (-37.7--22) | -4.9 (-15.5-5.6) | -20.5 (-28.8--12.2) | 2.0 (-8.4-12.4) |
| D1: 15-21 | 33.0 (30.1-35.9) | 72.2 (67.2-77.3) | 25.2 (20.4-30.0) | 56.7 (50.1-63.2) | 15.9 (9.0-22.9) | 46.0 (37.8-54.2) | 15.9 (7.5-24.3) | 39.1 (29.3-49.0) |
| D1: 22-28 | 47.0 (44.3-49.8) | 83.0 (78.8-87.1) | 45.8 (41.6-50.0) | 80.6 (76.3-84.9) | 42.9 (37.4-48.5) | 68.6 (62.6-74.6) | 23.4 (14.6-32.2) | 52.3 (43.2-61.5) |
| D2: 0-7 | 51.0 (46.9-55.2) | 82.5 (75.4-89.5) | 56.6 (50.7-62.6) | 86.6 (81.1-92.2) | 49.3 (41.5-57.1) | 73.0 (64.8-81.2) | 55.6 (47.1-64.2) | 60.7 (49.0-72.5) |
| D2: 8-14 | 71.4 (69.0-73.8) | 80.2 (74.1-86.2) | 73.5 (69.8-77.3) | 82.7 (76.9-88.5) | 67.4 (62.2-72.5) | 82.3 (76.6-88.0) | 68.4 (62.8-74.0) | 85.0 (80.0-90.1) |
| D2: 15-30 | 81.1 (79.9-82.2) | 91.8 (89.4-94.2) | 83.8 (81.9-85.6) | 89.1 (85.9-92.3) | 86.4 (84.4-88.5) | 92.6 (90.2-94.9) | 84.7 (82.2-87.2) | 89.8 (86.7-92.8) |
| D2: 1M-2M | 75.9 (75.0-76.8) | 95.4 (94.4-96.5) | 75.6 (74.0-77.2) | 94.0 (92.6-95.4) | 82.5 (80.8-84.3) | 93.8 (92.3-95.3) | 82.3 (80.2-84.4) | 92.0 (90.2-93.8) |
| D2: 2M-3M | 71.4 (70.4-72.4) | 95.5 (94.5-96.5) | 70.1 (68.3-71.9) | 92.6 (91.2-94.1) | 71.7 (68.7-74.6) | 90.1 (87.8-92.5) | 75.9 (72.7-79.1) | 90.7 (88.4-93.1) |
| D2: 3M-4M | 68.4 (67.4-69.5) | 94.6 (93.6-95.7) | 62.8 (60.7-64.9) | 92.4 (91.0-93.8) | 63.4 (59.9-66.8) | 88.4 (86.1-90.8) | 60.9 (56.0-65.8) | 82.7 (78.9-86.6) |
| D2: 4M-5M | 60.3 (59.1-61.6) | 94.7 (93.7-95.7) | 57.1 (54.9-59.3) | 91.0 (89.5-92.5) | 52.3 (48.2-56.3) | 87.0 (84.6-89.4) | 38.2 (31.0-45.4) | 72.1 (66.6-77.5) |
| D2: 5M-6M | 59.3 (58.1-60.6) | 95.5 (94.7-96.3) | 52.5 (50.2-54.9) | 92.4 (91.2-93.5) | 44.1 (39.7-48.5) | 82.6 (79.9-85.4) | 30.4 (23.0-37.8) | 73.2 (68.3-78.1) |
| D2: > 6M | 58.6 (57.2-60.0) | 94.0 (92.9-95.2) | 52.8 (50.2-55.4) | 90.0 (88.3-91.6) | 36.9 (32.0-41.8) | 82.2 (79.5-85.0) | 27.4 (20.0-34.8) | 69.8 (64.8-74.8) |
| DR: 1-7 | 64.8 (62.0-67.6) | 95.7 (93.1-98.3) | 53.4 (49.0-57.9) | 93.1 (90.4-95.8) | 38.3 (28.9-47.6) | 85.8 (80.7-91.0) | 37.3 (21.5-53.0) | 62.8 (47.9-77.8) |
| DR > 7 | 93.0 (92.3-93.6) | 98.8 (98.0-99.6) | 92.5 (91.8-93.1) | 99.0 (98.7-99.4) | 89.9 (88.8-90.9) | 98.2 (97.7-98.7) | 89.0 (87.5-90.5) | 97.3 (96.5-98.1) |

**Table 4.**
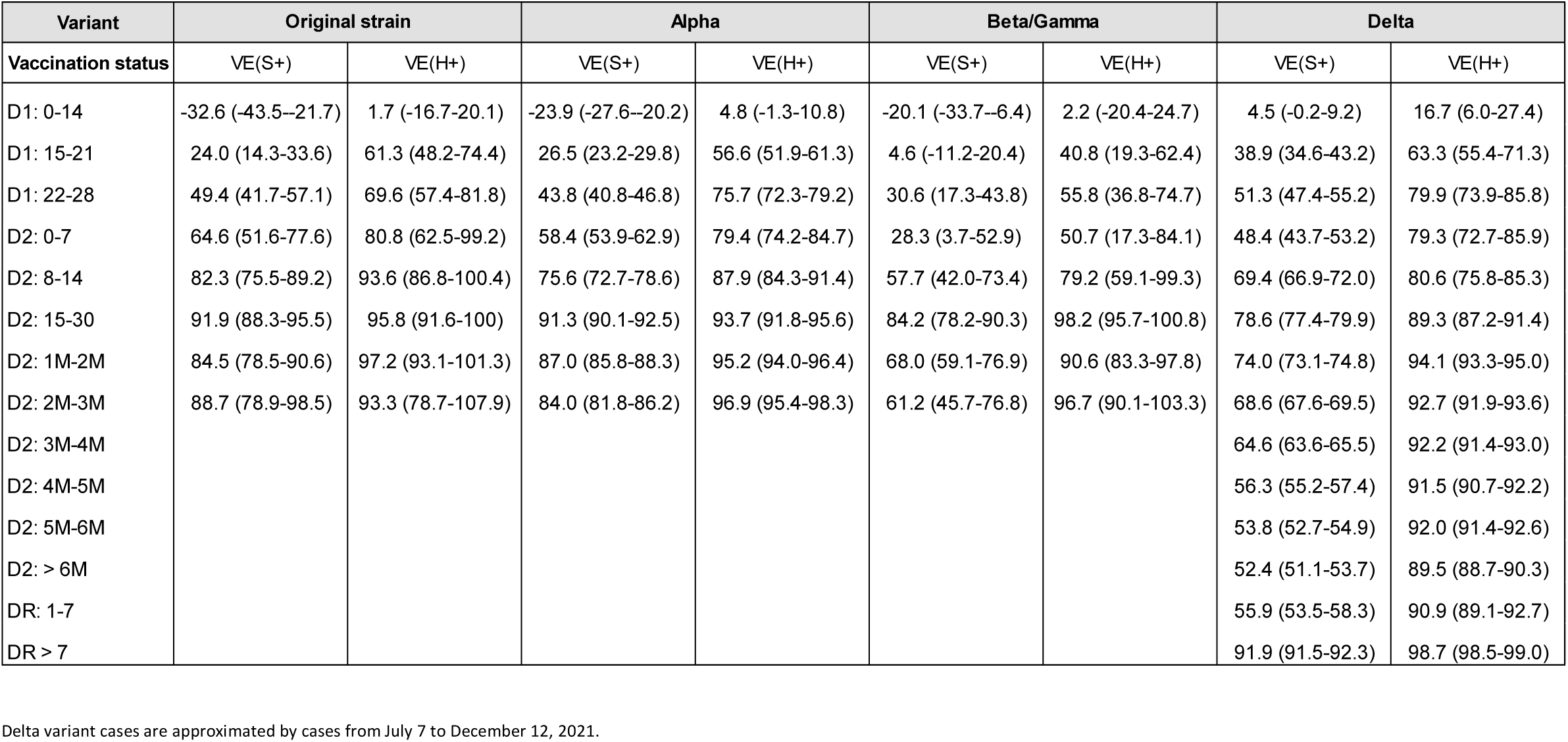
Covid-19 vaccine effectiveness (in %) by variant of concern against symptomatic infections and hospitalizations among persons aged 50 years or over, according to the time elapsed since the injection of each vaccine dose, data collected from January 1^st^ to December 12, 2021.

| Variant | Original strain |  | Alpha |  | Beta/Gamma |  | Delta |  |
| --- | --- | --- | --- | --- | --- | --- | --- | --- |
| Vaccination status | VE(S+) | VE(H+) | VE(S+) | VE(H+) | VE(S+) | VE(H+) | VE(S+) | VE(H+) |
| D1: 0-14 | -32.6 (-43.5--21.7) | 1.7 (-16.7-20.1) | -23.9 (-27.6--20.2) | 4.8 (-1.3-10.8) | -20.1 (-33.7--6.4) | 2.2 (-20.4-24.7) | 4.5 (-0.2-9.2) | 16.7 (6.0-27.4) |
| D1: 15-21 | 24.0 (14.3-33.6) | 61.3 (48.2-74.4) | 26.5 (23.2-29.8) | 56.6 (51.9-61.3) | 4.6 (-11.2-20.4) | 40.8 (19.3-62.4) | 38.9 (34.6-43.2) | 63.3 (55.4-71.3) |
| D1: 22-28 | 49.4 (41.7-57.1) | 69.6 (57.4-81.8) | 43.8 (40.8-46.8) | 75.7 (72.3-79.2) | 30.6 (17.3-43.8) | 55.8 (36.8-74.7) | 51.3 (47.4-55.2) | 79.9 (73.9-85.8) |
| D2: 0-7 | 64.6 (51.6-77.6) | 80.8 (62.5-99.2) | 58.4 (53.9-62.9) | 79.4 (74.2-84.7) | 28.3 (3.7-52.9) | 50.7 (17.3-84.1) | 48.4 (43.7-53.2) | 79.3 (72.7-85.9) |
| D2: 8-14 | 82.3 (75.5-89.2) | 93.6 (86.8-100.4) | 75.6 (72.7-78.6) | 87.9 (84.3-91.4) | 57.7 (42.0-73.4) | 79.2 (59.1-99.3) | 69.4 (66.9-72.0) | 80.6 (75.8-85.3) |
| D2: 15-30 | 91.9 (88.3-95.5) | 95.8 (91.6-100) | 91.3 (90.1-92.5) | 93.7 (91.8-95.6) | 84.2 (78.2-90.3) | 98.2 (95.7-100.8) | 78.6 (77.4-79.9) | 89.3 (87.2-91.4) |
| D2: 1M-2M | 84.5 (78.5-90.6) | 97.2 (93.1-101.3) | 87.0 (85.8-88.3) | 95.2 (94.0-96.4) | 68.0 (59.1-76.9) | 90.6 (83.3-97.8) | 74.0 (73.1-74.8) | 94.1 (93.3-95.0) |
| D2: 2M-3M | 88.7 (78.9-98.5) | 93.3 (78.7-107.9) | 84.0 (81.8-86.2) | 96.9 (95.4-98.3) | 61.2 (45.7-76.8) | 96.7 (90.1-103.3) | 68.6 (67.6-69.5) | 92.7 (91.9-93.6) |
| D2: 3M-4M |  |  |  |  |  |  | 64.6 (63.6-65.5) | 92.2 (91.4-93.0) |
| D2: 4M-5M |  |  |  |  |  |  | 56.3 (55.2-57.4) | 91.5 (90.7-92.2) |
| D2: 5M-6M |  |  |  |  |  |  | 53.8 (52.7-54.9) | 92.0 (91.4-92.6) |
| D2: > 6M |  |  |  |  |  |  | 52.4 (51.1-53.7) | 89.5 (88.7-90.3) |
| DR: 1-7 |  |  |  |  |  |  | 55.9 (53.5-58.3) | 90.9 (89.1-92.7) |
| DR > 7 |  |  |  |  |  |  | 91.9 (91.5-92.3) | 98.7 (98.5-99.0) |

635 Delta variant cases are approximated by cases from July 7 to December 12, 2021.

## Appendix 4 Sensitivity analyses

Presence of comorbidities is derived either from prior knowledge from the health insurance on the person status or because a physician recommended to prioritize a non-eligible (at-the-time) individual during the vaccination campaign. Thus, the latter is rare for unvaccinated persons. An alternative specification excludes from the sample persons who are classified as comorbid on a physician recommendation in order to benefit from the vaccine in priority. In this alternative specification, the odd ratio in the early period post first injection indicates a decreased risk [OR: 0.93 (CI 95% 91-95)] instead of the increase risk observed in the baseline specification. The comparison of our estimates to this alternative may inform on the extent of the downward bias due to targeting individuals on unobserved factors. The downward bias in the vaccine effectiveness against symptomatic diseases is very limited in the first month following the second injection, but could reach six percentage points after four months. In contrast, vaccine effectiveness against hospitalization are very similar in both alternatives.

**Figure 7.**
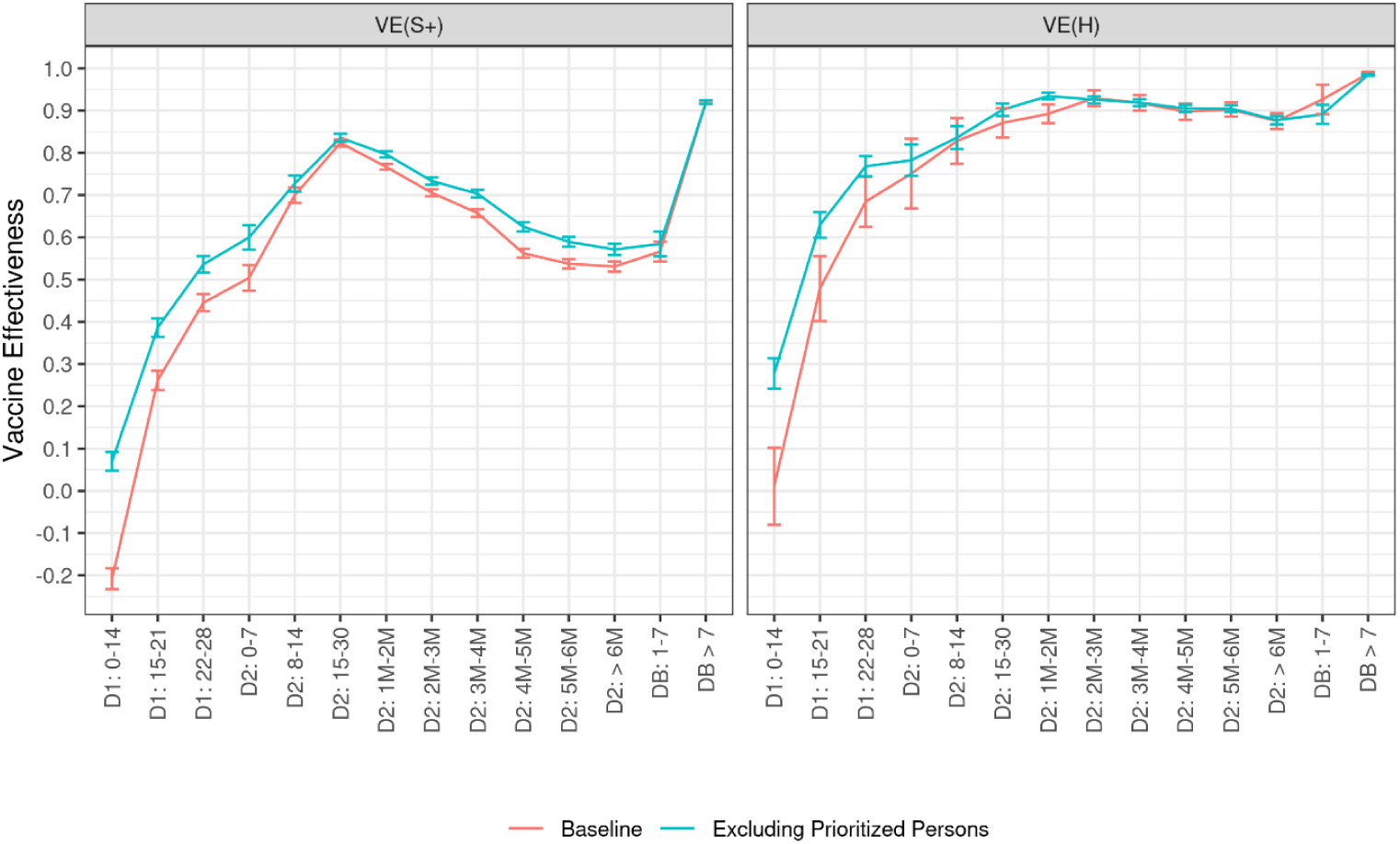
Covid-19 vaccine effectiveness against symptomatic infections and hospitalizations among persons aged 50 years or over, according to the time elapsed since the injection of each vaccine dose, data collected from January 1^st^ to December 12, 2021, when excluding persons prioritized by a physician for vaccine administration.

## Appendix 5 Matching characteristics

Among persons aged 50 years or over hospitalized for Covid-19 (data source: SI-VIC), 72 % have a matched positive RT-PCR test, collected from fifteen days before admission to the end of their stay (source: SI-DEP) over our analysis period. Among persons aged 50 years or over hospitalized for Covid-19 (data source: SI-VIC), 73 % have a match in the VAC-SI register, which covers nearly all French residents and allows to recover the vaccination status. Among persons aged 50 years reporting symptoms in the last seven days before a RT-PCR test (data source: SI-DEP), 84 % have a match in the VAC-SI register, thus a known vaccination status.

## References

Andrews, N., Tessier, E., Stowe, J., Gower, C., Kirsebom, F., Simmons, R., Gallagher, E., Chand, M., Brown, K., Ladhani, S. and Ramsay, M., (2021a). Vaccine effectiveness and duration of protection of Comirnaty, Vaxzevria and Spikevax against mild and severe COVID-19 in the UK. medRxiv.

Andrews, N., Stowe, J., Kirsebom, F., Gower, C., Ramsay, M. and Bernal, J.L., (2021b). Effectiveness of BNT162b2 (Comirnaty, Pfizer-BioNTech) COVID-19 booster vaccine against covid-19 related symptoms in England: test negative case-control study. medRxiv.

Andrews, Nick, et al. “Effectiveness of COVID-19 vaccines against the Omicron (B. 1.1. 529) variant of concern.” MedRxiv (2021c).

Bar-On, Y.M., Goldberg, Y., Mandel, M., Bodenheimer, O., Freedman, L., Kalkstein, N., Mizrahi, B., Alroy-Preis, S., Ash, N., Milo, R. and Huppert, A. (2021). Protection of BNT162b2 vaccine booster against Covid-19 in Israel. New England Journal of Medicine, 385(15), pp. 1393–1400.

Bouillon, K., Baricault, B., Botton, J., Jabagi, M.J., Bertrand, M., Semenzato, L., Le Vu, S., Drouin, J., Dray-Spira, R., Weill, A., Zureik, M. (2021, octobre). Estimation de l’impact de la vaccination chez les personnes âgées de 75 ans et plus sur le risque de formes graves de Covid-19 en France à partir des données du Système national des données de santé (SNDS) – actualisation jusqu’au 20 juillet 2021. Rapport d’études EPI-PHARE-Groupement d’intérêt scientifique (GIS) ANSM-CNAM.

Botton J, Dray-Spira R, Baricault B, Drouin J, Bertrand M, Jabagi MJ, Weill A, Zureik M. Reduced risk of severe COVID-19 in more than 1.4 million elderly people aged 75 years and older vaccinated with mRNA-based vaccines. Vaccine. 2022 Jan 24;40(3):414–7.

Buchan, Sarah A., et al. “Effectiveness of COVID-19 vaccines against Omicron or Delta infection.” medRxiv (2022): 2021–12.

Collie, Shirley, et al. “Effectiveness of BNT162b2 vaccine against omicron variant in South Africa.” New England Journal of Medicine (2021).

Chung, H., He, S., Nasreen, S., Sundaram, M.E., Buchan, S.A., Wilson, S.E., Chen, B., Calzavara, A., Fell, D.B., Austin, P.C. and Wilson, K.(2021). Effectiveness of BNT162b2 and mRNA-1273 covid-19 vaccines against symptomatic SARS-CoV-2 infection and severe covid-19 outcomes in Ontario, Canada: test negative design study, BMJ 374

Dean, N.E., Hogan, J.W. and Schnitzer, M.E. (2021). Covid-19 Vaccine Effectiveness and the Test-Negative Design. The New England Journal of Medicine.

Drees (2021). Les appariements SI-VIC, SI-DEP et VAC-SI

Goel, R.R., Painter, M.M., Apostolidis, S.A., Mathew, D., Meng, W., Rosenfeld, A.M., Lundgreen, K.A., Reynaldi, A., Khoury, D.S., Pattekar, A. and Gouma, S. (2021). mRNA Vaccination induces durable immune memory to SARS-CoV-2 with continued evolution to variants of concern. Biorxiv.

Goldberg, Y., Mandel, M., Bar-On, Y.M., Bodenheimer, O., Freedman, L., Haas, E.J., Milo, R., Alroy-Preis, S., Ash, N. and Huppert, A. (2021). Waning immunity after the BNT162b2 vaccine in Israel. New England Journal of Medicine.

Ireland, G., Whitaker, H., Shamez N Ladhani, S.N., Baawuah F., Subbarao, S., Linley E., Warrener, L., O’Brien, M., Whillock, C., Martin, O., Moss, P., Ramsay, M.E., Amirthalingam, G., Brown, K.E. (2022). Serological responses to COVID-19 Comirnaty booster vaccine, London, United Kingdom, September to December 2021. Eurosurveillance.

Jabagi, M.J., Botton, J., Baricault, B., Bouillon, K., Bertrand, M., Semenzato, L., Le Vu, S., Drouin, J., Weill, A., Dray-Spira, R., Zureik, M. (2021, octobre). Estimation de l’impact de la vaccination sur le risque de formes graves de Covid-19 chez les personnes de 50 à 74 ans en France à partir des données du Système national des données de santé (SNDS). Rapport d’études EPI-PHARE - Groupement d’intérêt scientifique (GIS) ANSM-CNAM.

Jackson, M.L. and Nelson, J.C., (2013). The Test-Negative Design for estimating influenza vaccine effectiveness. Vaccine, 31(17), pp. 2165–2168.

Breslow, N.E., Day, N.E., Halvorsen, K.T., Prentice, R.L. and Sabai, C., (1978). American Journal of Epidemiology, 108(4), pp. 299–307.

Levin, E.G., Lustig, Y., Cohen, C., Fluss, R., Indenbaum, V., Amit, S., Doolman, R., Asraf, K., Mendelson, E., Ziv, A. and Rubin, C. (2021). Waning immune humoral response to BNT162b2 Covid-19 vaccine over 6 months. New England Journal of Medicine.

Lopez Bernal J, Andrews N, Gower C, Robertson C, Stowe J, Tessier E, et al. (2021). Effectiveness of the Pfizer-BioNTech and Oxford-AstraZeneca vaccines on covid-19 related symptoms, hospital admissions, and mortality in older adults in England: test negative case-control study. BMJ, 373.

Lopez Bernal, J., Andrews, N., Gower, C., Gallagher, E., Simmons, R., Thelwall, S., Stowe, J., Tessier, E., Groves, N., Dabrera, G. and Myers, R. (2021). Effectiveness of Covid-19 vaccines against the B. 1.617. 2 (Delta) variant. N Engl J Med, pp. 585–594.

Mattiuzzi, C., Lippi, G. (2022). Primary COVID-19 vaccine cycle and booster doses efficacy: analysis of Italian nationwide vaccination campaign. European Journal of Public Health.

Santé Publique France (2021, juillet). Encourager la vaccination face à l’augmentation de la circulation variants : un enjeu majeur pour continuer à contenir l’épidémie. Point épidémiologique Covid-19 du 1er juillet 2021

Stowe, J., Andrews, N., Gower, C., Gallagher, E., Utsi, L. and Simmons, R. (2021). Effectiveness of COVID-19 vaccines against hospital admission with the Delta (B. 1.617. 2) variant. Public Health England.

Tartof, S.Y., Slezak, J.M., Fischer, H., Hong, V., Ackerson, B.K., Ranasinghe, O.N., Frankland, T.B., Ogun, O.A., Zamparo, J.M., Gray, S. and Valluri, S.R. (2021). Effectiveness of mRNA BNT162b2 COVID-19 vaccine up to 6 months in a large integrated health system in the USA: a retrospective cohort study. The Lancet, 398(10309), pp. 1407–1416.

Tenforde, M.W., Patel, M.M., Ginde, A.A., Douin, D.J., Talbot, H.K., Casey, J.D., Mohr, N.M., Zepeski, A., Gaglani, M., McNeal, T. and Ghamande, S. (2021). Effectiveness of SARS-CoV-2 mRNA vaccines for preventing Covid-19 hospitalizations in the United States. Clin. Infect. Dis.

Thomas, Stephen J., et al. “Safety and efficacy of the BNT162b2 mRNA Covid-19 vaccine through 6 months.” New England Journal of Medicine 385.19 (2021): 1761–1773.

Thompson, M.G., Stenehjem, E., Grannis, S., Ball, S.W., Naleway, A.L., Ong, T.C., DeSilva, M.B., Natarajan, K., Bozio, C.H., Lewis, N. and Dascomb, K. (2021). Effectiveness of Covid-19 vaccines in ambulatory and inpatient care settings. New England Journal of Medicine, 385(15), pp. 1355–1371.

Willett, Brian J., et al. “The hyper-transmissible SARS-CoV-2 Omicron variant exhibits significant antigenic change, vaccine escape and a switch in cell entry mechanism.” medRxiv (2022).

World Health Organization (2021). Evaluation of COVID-19 vaccine effectiveness: interim guidance, 17 March 2021 (No. WHO/2019-nCoV/vaccine_effectiveness/measurement/2021.1).

